# Distinguishing GPT-4-generated Radiology Abstracts from Original Abstracts: Performance of Blinded Human Observers and AI Content Detector

**DOI:** 10.1101/2023.04.28.23289283

**Authors:** Furkan Ufuk, Hakki Peker, Ergin Sagtas, Ahmet Baki Yagci

## Abstract

**Objective:** To determine GPT-4’s effectiveness in writing scientific radiology article abstracts and investigate human reviewers’ and AI Content detectors’ success in distinguishing these abstracts. Additionally, to determine the similarity scores of abstracts generated by GPT-4 to better understand its ability to create unique text.

**Methods:** The study collected 250 original articles published between 2021 and 2023 in five radiology journals. The articles were randomly selected, and their abstracts were generated by GPT-4 using a specific prompt. Three experienced academic radiologists independently evaluated the GPT-4 generated and original abstracts to distinguish them as original or generated by GPT-4. All abstracts were also uploaded to an AI Content Detector and plagiarism detector to calculate similarity scores. Statistical analysis was performed to determine discrimination performance and similarity scores.

**Results:** Out of 134 GPT-4 generated abstracts, average of 75 (56%) were detected by reviewers, and average of 50 (43%) original abstracts were falsely categorized as GPT-4 generated abstracts by reviewers. The sensitivity, specificity, accuracy, PPV, and NPV of observers in distinguishing GPT-4 written abstracts ranged from 51.5% to 55.6%, 56.1% to 70%, 54.8% to 60.8%, 41.2% to 76.7%, and 47% to 62.7%, respectively. No significant difference was observed between observers in discrimination performance.

**Conclusion:** GPT-4 can generate convincing scientific radiology article abstracts. However, human reviewers and AI Content detectors have difficulty in distinguishing GPT-4 generated abstracts from original ones.

## Introduction

Recent progress in natural language processing has led to the development of large language models (such as GPT-4) capable of generating high-quality text. GPT-4 has gained widespread attention due to its ability to generate coherent and informative text on various topics (1-4). However, there is a need to determine its ability to write convincing medical research articles. Moreover, while the similarity score of all published original abstracts is expected to be complete, the similarity scores of the abstracts created by GPT-4 are unknown. We aimed to evaluate the effectiveness of GPT-4 in writing scientific radiology article abstracts and investigate human reviewers’ and AI Content detectors’ success in distinguishing these abstracts. We also aimed to determine the similarity scores of abstracts generated by GPT-4 to better understand the ability of GPT-4 to create unique text.

## Methods

A radiology resident (*Reviewer*) collected a total of 250 original articles that were published between 2021 and 2023 in the *Radiology, European Radiology, American Journal of Roentgenology, Diagnostic and Interventional Radiology*, and *Japanese Journal of Radiology* were investigated. The articles are in five sub-specialties: Abdominal, Breast, Cardiothoracic, Musculoskeletal radiology, and Neuroradiology. The reviewer randomly selected 134 articles (53.6%) and uploaded the content to GPT-4, excluding the abstract section, and the abstracts were written by GPT-4.

The prompt fed to the GPT-4 was “Please write a scientific abstract for this article with a maximum word count of 300, using these same subheadings: Background, Objective, Methods, Results, and Conclusion.”. Then, the reviewer controlled and reformatted the subheadings of the abstract according to the specific style and requirements of the journal, as follows:

1. *Radiology:* Background, Purpose, Materials and Methods, Results, and Conclusion.
2. *European Radiology:* Objective, Materials and methods, Results, Conclusions.
3. *American Journal of Roentgenology:* Background, Objective, Methods, Results, and Conclusion.
4. *Diagnostic and Interventional Radiology:* PURPOSE, METHODS, RESULTS, and CONCLUSION.
5. *Japanese Journal of Radiology:* Purpose, Materials and Methods, Results, and Conclusion.

Three experienced academic radiologists with 8 (F.U., cardiothoracic radiologist), 21 (E.S., neuroradiologist), and 22 (A.B.Y., urogenital radiologist) years of experience independently evaluated the GPT-4 generated and the original abstracts to distinguish them as original or generated by GPT-4. Observers were utterly unaware of how many abstracts were written by ChatGPT, and the observers were blinded to the source of each abstract. Although journals had their specific abstract formats, it was not among the observers’ aim to distinguish which journal the abstracts belonged to.

The abstracts were uploaded to the widely used plagiarism detector (https://www.ithenticate.com), and similarity scores were calculated. iThenticate uses advanced algorithms and techniques to compare the text of a given document with a vast database of scholarly publications, internet sources, and other documents to identify any instances of potential plagiarism or text reuse. It provides users with a similarity report that highlights any matches found and allows them to review and verify the originality of their work.

Moreover, all abstracts were uploaded to artificial intelligence (AI) Content Detector (https://contentatscale.ai), and “Human Content Scores” were calculated. “Human Content Score” in this context refers to a metric or rating that reflects the quality, relevance, or engagement level of written content as evaluated by human experts.

Observers’ ability to discriminate was investigated using diagnostic tests and a two-sample t-test used to determine if there was a significant difference between the observers in discrimination performance. The receiver operating characteristic (ROC) analysis and area under the ROC curve (AUROC) were used to assess the discriminating performance of the human content score for GPT-4 and original abstracts.

Statistical analyses were performed using MedCalc version 20 (MedCalc Software) and SPSS version 23 (IBM). A P value *<.05* was considered statistically significant.

## Results

Seventy-five of the 134 (56%) GPT-4 generated abstracts were detected by reviewers on average. Fifty of the 116 (43%) original abstracts were falsely categorized as GPT-4 generated abstracts by reviewers on average. The sensitivity, specificity, accuracy, positive predictive value (PPV), and negative predictive value (NPV) of observers in distinguishing GPT-4 written abstracts ranged from 51.5–55.6%, 56.1–70%, 54.8–60.8%, 41.2–76.7%, and 47–62.7%, respectively (Table 1). There was no significant difference between observers in discrimination performance (*P =.074* for Observer 1 and Observer 2, *P =.521* for Observer 1 and Observer 3, and *P =.233* for Observer 2 and Observer 3).

**Table 1:** Performance of blinded human observers to distinguish GPT-4-generated abstracts from the original abstracts.

|  |  | Original<br>(n) | GPT-4<br>(n) | Sens.<br>(%) | Spec.<br>(%) | Acc.<br>(%) | PPV<br>(%) | NPV<br>(%) |
| --- | --- | --- | --- | --- | --- | --- | --- | --- |
| *Observer 1* | *Original* | 53 | 63 | 51.5 | 57.1 | 54.8 | 41.2 | 62.7 |
|  | *GPT-4* | 50 | 84 |  |  |  |  |  |
| *Observer 2* | *Original* | 89 | 27 | 55.6 | 70 | 60.8 | 76.7 | 47 |
|  | *GPT-4* | 71 | 63 |  |  |  |  |  |
| *Observer 3* | *Original* | 65 | 61 | 53.7 | 56.1 | 57.2 | 51.6 | 58.2 |
|  | *GPT-4* | 56 | 78 |  |  |  |  |  |

Sens., sensitivity; Spec., specificity; Acc., accuracy; PPV, positive predictive value; NPV, negative predictive value; n, number of cases.

While Observer 2 and 3 performed best in discriminating GPT-4 generated abstracts in *Radiology*, Observer 1 showed in the *American Journal of Roentgenology*. However, according to the journal, there was no significant difference in the observers’ ability to discriminate (*P =.057* for Observer 1, *P =.275* for Observer 2, and *P =.051* for Observer 3). Moreover, no statistically significant difference was found between the author’s discrimination performance in different subspecialties (*P ≥.243*) (Fig. 1).

**Figure 1:**
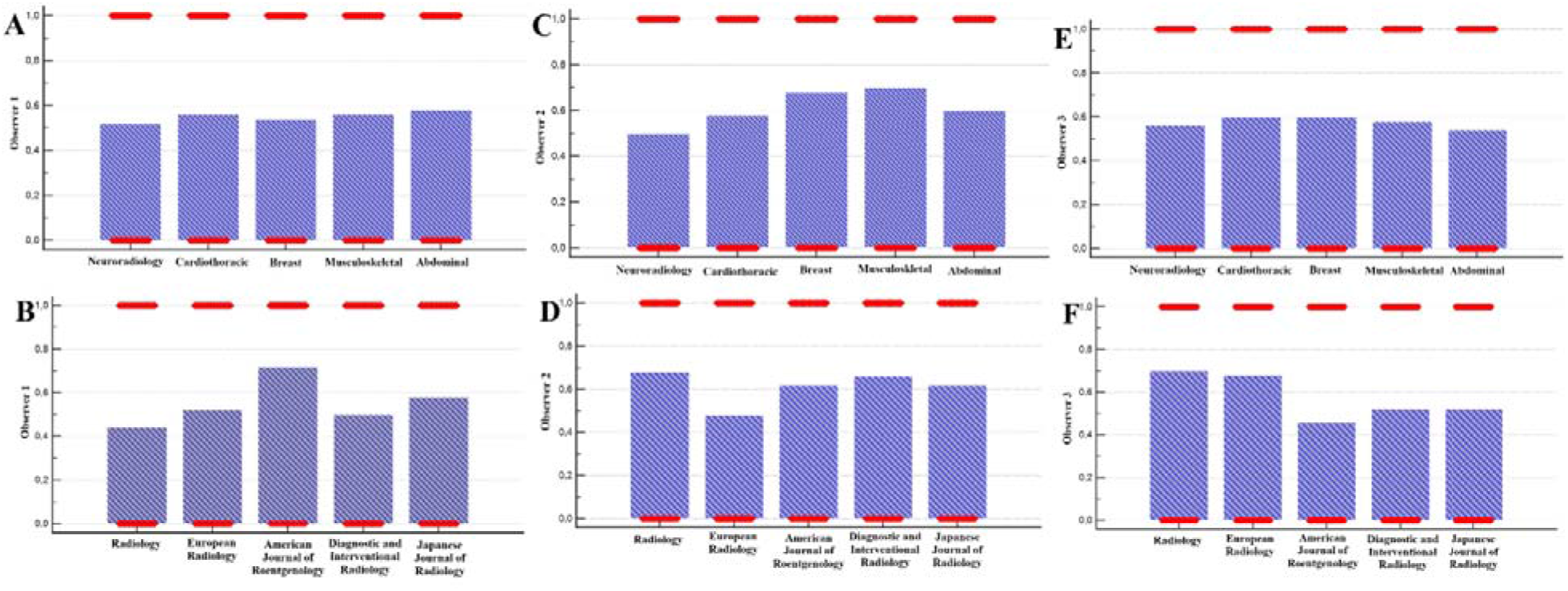
Accuracies of observers in distinguishing GPT-4-generated abstracts from originals according to the journal and subspecialty. Observer’s accuracies according to the (**A, C, E)** Subspecialties and **(B, D, F)** Journals.

The mean plagiarism (similarity) score of the abstracts generated by GPT-4 was 57.6 ± 12.3%. The discriminative value of the plagiarism detector was not affected by the journal or subspecialty (*P =.101* and *P =.543*, respectively) (Fig. S1).

The median human content score for GPT-4 abstracts was 90% (range, 48–100%) and 91% (range, 44–100%) for original abstracts. The AUROC value of the human content score was 0.564 (95% confidence interval, 0.499–0.626), and a cutoff value of <94 indicated GPT-4 generated abstract with a 73% sensitivity and 40.6% specificity. No significant difference in the AI-content detector score according to the journal or subspecialty (*P =.608* and *P =.091*, respectively) (Fig. S2).

## Discussion

This study found that AI-content and the plagiarism detectors and human observers’ performance in distinguishing GPT-4 written abstracts was limited. No significant effect of journal and subspecialty was detected on distinguishability. These findings suggest that GPT-4 has the potential to write scientific article abstracts in radiology effectively.

Blinded human observers’ low accuracy in detecting GPT-4-generated abstracts may be due to the algorithm’s ability to produce text that resembles human-written text. Similarly, Gao et al. (5) showed that human observers detected only 68% of ChatGPT-generated scientific abstracts correctly.

Using a single plagiarism detection and AI-content detector and the lack of standardization in abstract reviewing processes are limitations of this study. However, despite its limitations, the study’s relevance to radiology, the random selection of articles from various radiology journals with observer blinding, and the evaluation of abstracts by experienced academic radiologists add credibility to the study’s findings.

The results of this study provide valuable insights into the potential of GPT-4 in radiology article writing and its implications for scientific publishing.

## Data Availability

All data produced in the present study are available upon reasonable request to the authors

## Supplementary Figures

**Figure S1:**
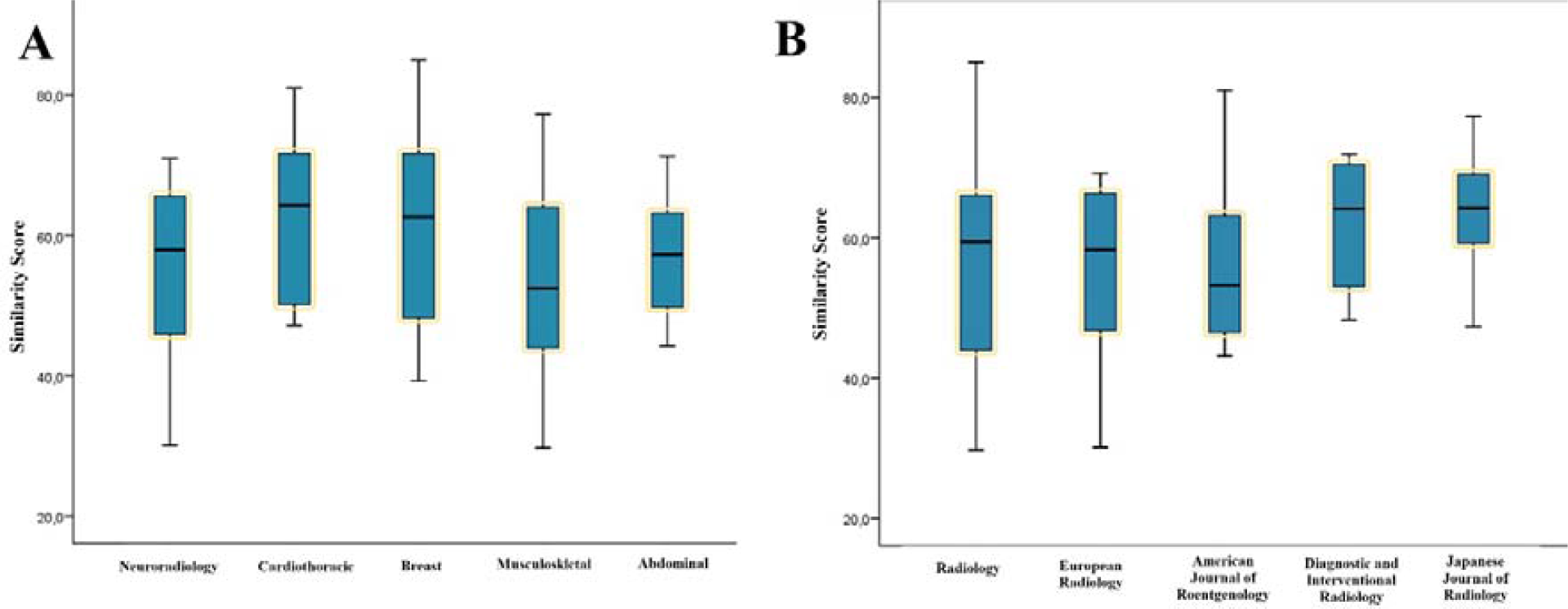
Plagiarism detection scores in GPT-4-generated abstracts according to the **(A)** Subspecialty and **(B)** Journal.

**Figure S2:**
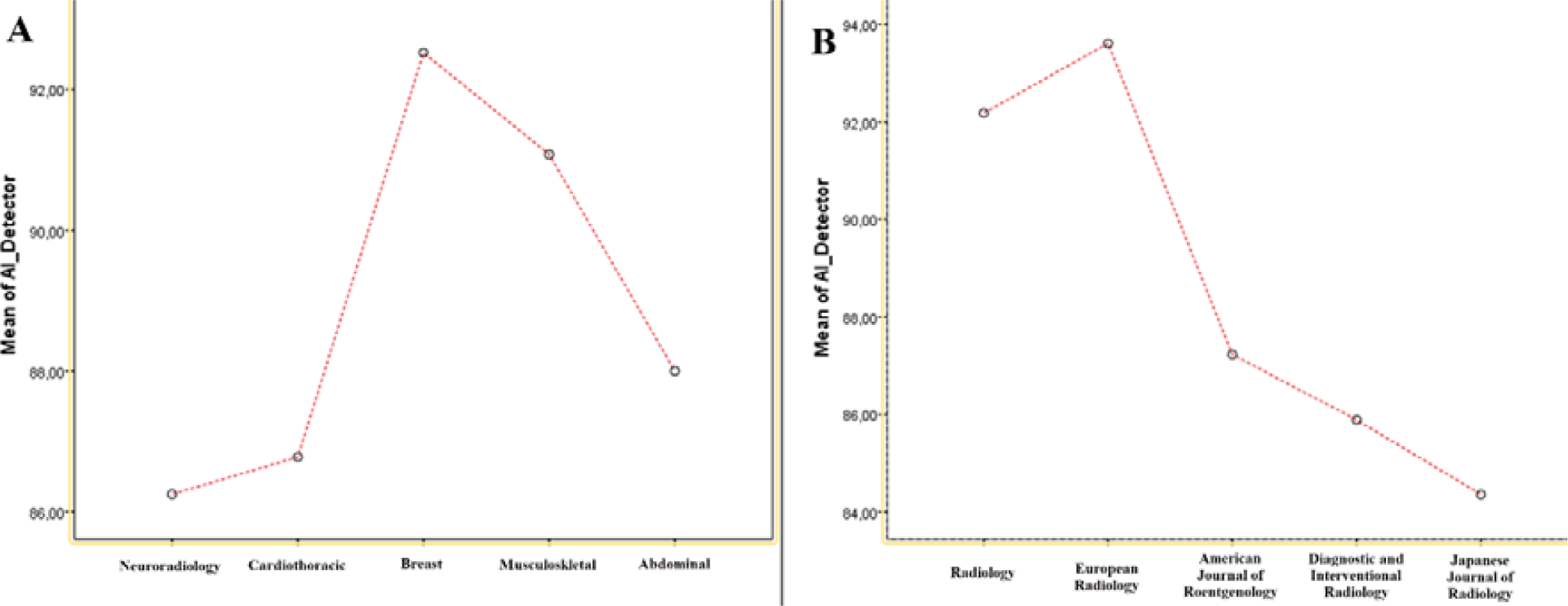
AI-content detector scores according to the **(A)** Subspecialty and **(B)** Journal.

